# Knowledge, Attitudes, and Practices Regarding Urinary Tract Infections Among Women in the United Arab Emirates

**DOI:** 10.1101/2024.02.05.24302366

**Authors:** Shaikha Salah Alhaj, Sajad Allami, Amjad Mohamadiyeh, Ammar Agha, Abdul Kareem Abu Ali, Joudi M Bassam Habbal, Balsam Qubais Saeed

**Author notes:** **Corresponding Author:** Shaikha Salah Alhaj, College of Medicine, University of Sharjah, Sharjah, UAE.

## Abstract

**Background:** Urinary tract infections (UTIs), which are infections of the kidneys, ureters, bladder, or urethra, are a worldwide public health concern. As compared to men, women are more prone to UTIs. There have been several studies that explore the knowledge, attitudes, and practices of women regarding UTIs in different countries, but no such study has been conducted in the UAE; therefore, we conducted this study in the UAE setting.

**Methods:** This study was conducted using an online survey created on Microsoft Forms. The minimum sample size required for our study was 385. This study was conducted after obtaining ethical approval from the Research Ethics Committee at the University of Sharjah (Reference Number: REC-23-09-19-01-F). A personally designed questionnaire consisting of 21 items, derived from previous research was used to record data. The data was analyzed using SPSS.

**Results:** A total of 475 females were included in the study. Most respondents were aged 18-30 years (47.4%). Our study found that a majority of the participants (69.7%) correctly identified bacteria as the most common cause of UTIs. With regards to practices undertaken during UTIs, among the participants with a history of UTI episodes, 32.6% waited 24-48 hours before seeking medical attention at a hospital or clinic, while 10% did not visit a hospital at all. Distinct trends were found when comparing demographic factors with knowledge levels. Most notably, the age group of 18-30 years showed the highest percentage of high-knowledge individuals (49%) compared to other age groups (p < 0.05). Education level was significantly (p = 0.003) associated with UTI knowledge. Going to the hospital/clinic was reported by 41% with high knowledge but only by 20% of those with poor knowledge. Moreover, a higher proportion of individuals with high knowledge sought medical attention immediately within 24 hours (47%).

**Conclusion:** Most of the participants possessed adequate knowledge regarding UTIs. Higher knowledge levels were associated with more proactive & appropriate health behaviors, such as seeking medical attention promptly & drinking more water.

## Introduction

Urinary tract infections (UTIs), which are infections of the kidneys, ureters, bladder, or urethra, are a worldwide public health concern [1]. According to a study, over 404.6 million people all around the globe had UTIs in 2019, and nearly 236,786 died as a result of UTIs the same year [2]. As compared to men, women are more prone to UTIs [1]. One of the reasons attributable to this fact is the presence of shorter urethra in women, which makes it easier for bacteria or other microbes to reach the bladder or urinary tract and cause infection [1, 3]. It is estimated that approximately 60% of females experience at least one UTI in their lifetime [3], which is more likely to occur in menopausal or pregnant women [3-4]. The urinary system is designed to keep the foreign bacteria out, but sometimes, failure of host defenses to remove these bacteria can result in UTIs. *Escherichia coli* is the most associated bacteria with UTIs. Other causative pathogens include *Staphylococcus saprophyticus, Proteus, Klebsiella*, and *Pseudomonas aeruginosa* [5-6]. Additionally, certain risk factors predispose women to UTIs, which include pregnancy, sexual activity (new sexual partner), menopause, use of certain birth controls, catheters, or a suppressed immune system [5-6]. UTIs can be both symptomatic and asymptomatic. Symptomatic UTIs are characterized by painful and frequent urination, hematuria, suprapubic pain, and fever [4-5]. On the other hand, asymptomatic UTIs do not have any apparent symptoms but are characterized by the presence of bacteria in urine [5, 7]. However, both symptomatic and asymptomatic infections should be treated [7] to avoid further complications such as recurrent infections, kidney damage, bladder dysfunction, or preterm labor [6, 8].

Despite being preventable, UTIs are quite prevalent among women in the UAE [4]. They can be prevented by staying well-hydrated, maintaining hygiene, urinating after sexual activity, and raising awareness among girls and women [1, 6]. There have been several studies that explore the knowledge, attitudes, and practices of women regarding UTIs in different countries [9-10], but no such study has been conducted in the UAE; therefore, in this study, we aim to explore the knowledge, attitudes, and practices of women in the UAE regarding UTIs. The results of this study will aid us in determining the level of awareness among them, which can contribute to targeted awareness campaigns and educational initiatives related to women’s health.

## Methods & Materials

The Strengthening the Report of the Observational Studies in Epidemiology (STOBE) guidelines were used throughout conducting this study [11].

### Study Design

This study was a cross-sectional survey to assess the knowledge, attitude, and practices (KAP) regarding Urinary Tract Infections (UTI) among women in the United Arab Emirates (UAE).

### Study Settings and Selection of Participants

This study was conducted using an online survey created on Microsoft Forms. The minimum sample size required for our study was 385, collected using the Raosoft Sample Size Calculator with a confidence interval of 95%, a margin of error of 5%, and a population size of 3.2 million. The data for the population size of women in the UAE was used according to the latest population statistics [12]. However, we calculated data from 475 participants. The inclusion criteria for participants included: i) females of any age; ii) residents of the UAE. The exclusion criteria included: i) males; ii) not a resident of the UAE.

### Ethics Statement

Data was collected from participants after obtaining informed consent. The online consent was taken prior to filling out the questionnaire. A brief summary of the questionnaire ahead was provided along with the purpose of filling out this form. It was clarified that participation is anonymous and voluntary. Only those who consented to participate could access the form ahead. In the case of minors, it was mandatory that the consent form was filled out in the presence of parents or guardians. Participants were ensured that their information would be kept confidential. This study was conducted after obtaining ethical approval from the Research Ethics Committee at the University of Sharjah (Reference Number: REC-23-09-19-01-F).

### Data Collection and Statistical Analysis

A personally designed questionnaire consisting of 21 items, derived from previous research and expert opinion [9-10], was used to record data. It was divided into four sections: i) demographics details; ii) questions to determine knowledge of the respondents; iii) questions to determine the attitude of the respondents; iv) questions to determine practices of the respondents. Both English and Arabic versions of the questionnaire were used to collect data. Data collection was started on 24^th^ October 2023 and we completed collecting data on 1^st^ December 2023.

The data was collected and coded. It was then analyzed using IBM Statistical Package for Social Sciences (SPSS) for Windows, Version 21 (IBM Corp, Armonk, NY, USA). A descriptive analysis was run. A knowledge score was calculated, where a 0-1 score was categorized as poor knowledge, a 2-3 score was categorized as moderate knowledge, and a 4-6 score was categorized as high knowledge. To study the association of factors with knowledge, a chi-square test was run, and a p < 0.05 was used as the level of significance.

## Results

### 1. Characteristics of the Study Population

A total of 475 females were included in the study. Most respondents were aged 18-30 years (47.4%). The participants were equally distributed between married (48.8%) and single (48.2%), with a small minority divided between divorced and widowed. A significant majority of the participants held a bachelor’s degree (66.9%), followed by those with a high school diploma (18.1%). Most participants lived in Sharjah and Dubai (73.1%), while the rest were distributed among the remaining emirates. 35 participants (7.3%) had either diabetes or hypertension. A summary of the demographic characteristics of the participants is shown in ***Table 1***.

**Table 1:**
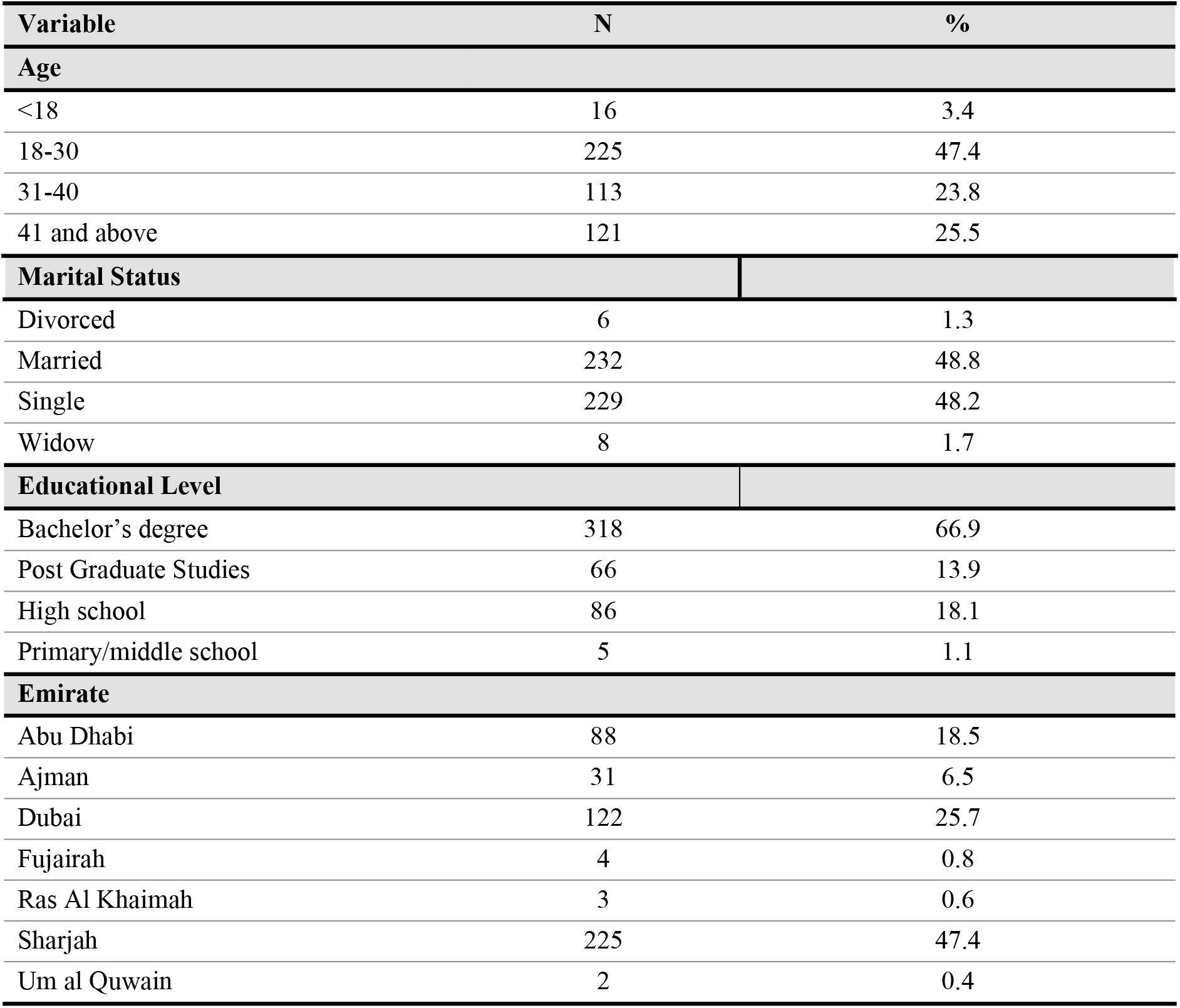
Demographic Characteristics of the Participants.

| Variable | N | % |
| --- | --- | --- |
| **Age** |  |  |
| <18 | 16 | 3.4 |
| 18-30 | 225 | 47.4 |
| 31-40 | 113 | 23.8 |
| 41 and above | 121 | 25.5 |
| **Marital Status** |  |  |
| Divorced | 6 | 1.3 |
| Married | 232 | 48.8 |
| Single | 229 | 48.2 |
| Widow | 8 | 1.7 |
| **Educational Level** |  |  |
| Bachelor's degree | 318 | 66.9 |
| Post Graduate Studies | 66 | 13.9 |
| High school | 86 | 18.1 |
| Primary/middle school | 5 | 1.1 |
| **Emirate** |  |  |
| Abu Dhabi | 88 | 18.5 |
| Ajman | 31 | 6.5 |
| Dubai | 122 | 25.7 |
| Fujairah | 4 | 0.8 |
| Ras Al Khaimah | 3 | 0.6 |
| Sharjah | 225 | 47.4 |
| Um al Quwain | 2 | 0.4 |

**Table 2:** Knowledge of the Participants Regarding Urinary Tract Infections.

| Variable | N | % |
| --- | --- | --- |
| **Have they heard of the term UTI?** |  |  |
| No | 89 | 18.7 |
| Yes | 386 | 81.3 |
| **Definition of UTI** |  |  |
| Inflammation of the bladder | 17 | 4.4 |
| Inflammation of the kidney | 2 | 0.5 |
| Inflammation of the urethra | 110 | 28.5 |
| None of the above | 3 | 0.8 |
| All of the above* | 254 | 65.8 |
| **Most common cause of UTI** |  |  |
| Bacteria* | 269 | 69.7 |
| Bad hygiene | 37 | 9.6 |
| Fungi | 58 | 15.0 |
| Viruses | 22 | 5.7 |
| **Which symptom/s do you think occur with UTI?** |  |  |
| Pain during urination* | 351 | 19.5 |
| Change in the urine color/appearance* | 302 | 16.8 |
| Abdominal pain* | 205 | 11.4 |
| Back pain* | 170 | 9.5 |
| Increase in the frequency of urination* | 251 | 14.0 |
| Leg pain | 31 | 1.7 |
| Fever* | 199 | 11.1 |
| Constipation | 22 | 1.2 |
| A sudden desire to go to bathroom to urinate* | 266 | 14.8 |
| **Which habit/s do you think increases the chances of having UTI?** |  |  |
| Cleaning the genitalia from back to front* | 238 | 27.5 |
| Drinking a small amount of water* | 334 | 38.6 |
| Delaying urination* | 267 | 30.8 |
| Urinating after sexual intercourse | 10 | 1.2 |
| Cleaning the genitalia from front to back | 9 | 1.0 |
| Drinking large amounts of water | 8 | 0.9 |
| **Which factor/s do you think prevent UTI?** |  |  |
| Cleaning the genitalia from back to front | 20 | 2.3 |
| Drinking a small amount of water | 3 | 0.3 |
| Delaying urination | 5 | 0.6 |
| Urinating after sexual intercourse* | 207 | 23.8 |
| Cleaning the genitalia from front to back* | 278 | 31.9 |
| Drinking large amounts of water* | 358 | 41.1 |

\* represents the correct answers

**Table 3:** Attitude of the Participants Regarding Urinary Tract Infections.

| Variable | N | % |
| --- | --- | --- |
| **In your opinion, what is the appropriate way of dealing with UTI?** |  |  |
| Drink more water | 74 | 19.2 |
| Go to the Hospital/Clinic | 269 | 69.6 |
| I don't know | 9 | 2.3 |
| Rest at home | 1 | 0.3 |
| Shower more frequently | 6 | 1.6 |
| Take analgesics (painkillers) | 1 | 0.3 |
| Take antibiotics directly | 26 | 6.7 |
| **Do you think UTIs are common?** |  |  |
| Yes | 363 | 94 |
| No | 23 | 6 |
| **Which of the following statements do you think is true regarding UTI?** |  |  |
| I don't know | 34 | 8.9 |
| UTIs affect both genders equally | 52 | 13.5 |
| UTIs affect females more | 295 | 76.3 |
| UTIs affect males more | 5 | 1.3 |
| **Which complications do you expect from having a UTI?** |  |  |
| It can damage the kidneys | 289 | 39.9 |
| It can decrease the quality of life | 171 | 23.6 |
| It can lead to recurrent UTIs | 142 | 19.6 |
| It can affect pregnancies | 85 | 11.8 |
| It can lead to death | 29 | 4 |
| It can cause weight loss | 8 | 1.1 |

| Do you feel UTI is a serious condition/disease? |  |  |
| --- | --- | --- |
| I don't know | 70 | 18.1 |
| Yes | 178 | 46.1 |
| No | 138 | 35.8 |

**Table 4:**
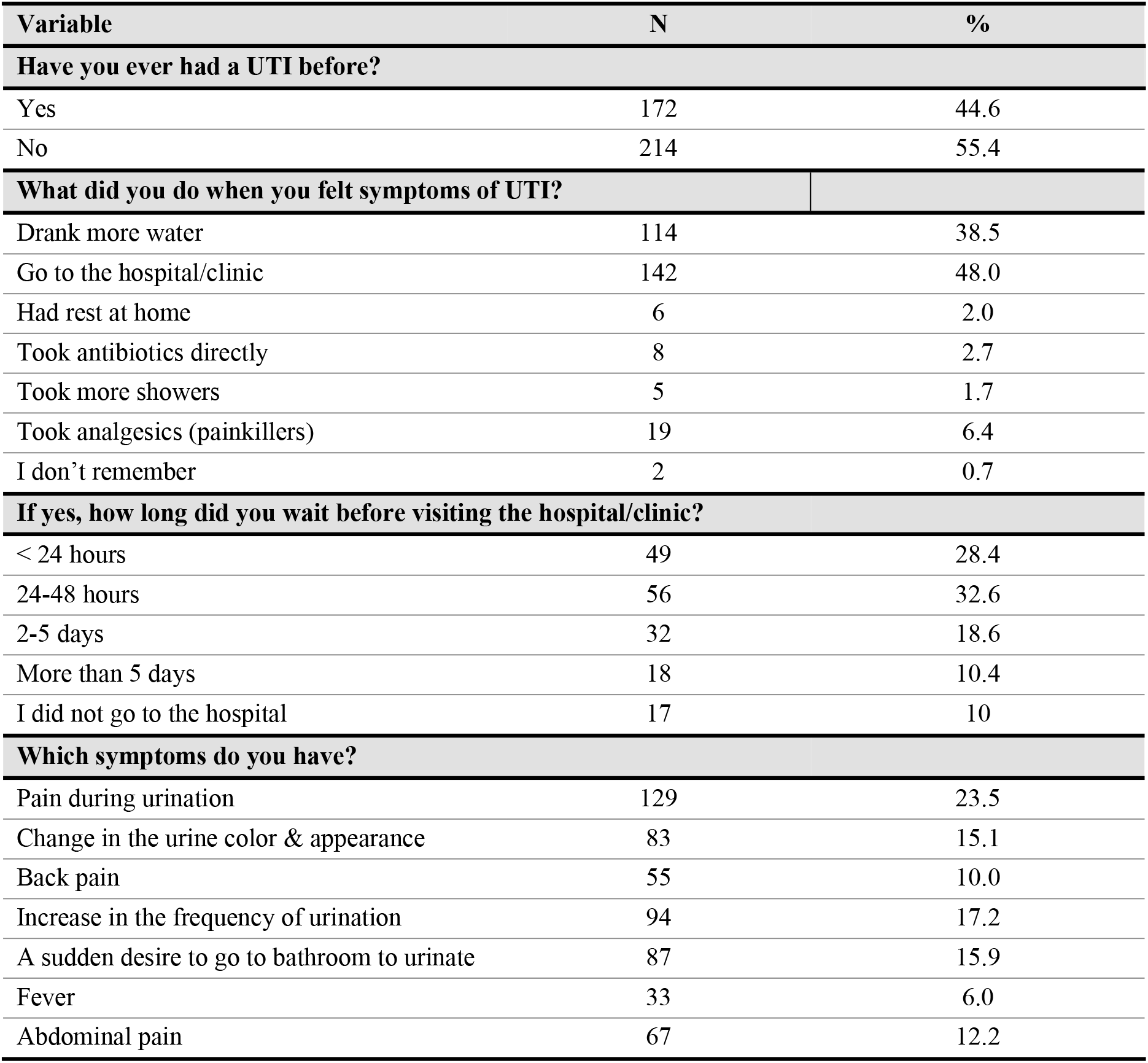
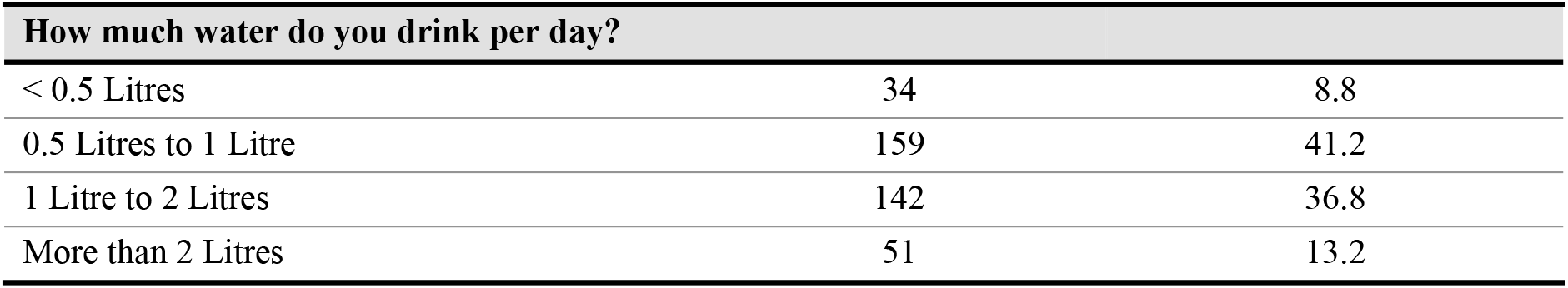
Practices of the Participants Regarding Urinary Tract Infections.

| Variable | N | % |
| --- | --- | --- |
| Have you ever had a UTI before? |  |  |
| Yes | 172 | 44.6 |
| No | 214 | 55.4 |
| What did you do when you felt symptoms of UTI? |  |  |
| Drank more water | 114 | 38.5 |
| Go to the hospital/clinic | 142 | 48.0 |
| Had rest at home | 6 | 2.0 |
| Took antibiotics directly | 8 | 2.7 |
| Took more showers | 5 | 1.7 |
| Took analgesics (painkillers) | 19 | 6.4 |
| I don't remember | 2 | 0.7 |
| If yes, how long did you wait before visiting the hospital/clinic? |  |  |
| < 24 hours | 49 | 28.4 |
| 24-48 hours | 56 | 32.6 |
| 2-5 days | 32 | 18.6 |
| More than 5 days | 18 | 10.4 |
| I did not go to the hospital | 17 | 10 |
| Which symptoms do you have? |  |  |
| Pain during urination | 129 | 23.5 |
| Change in the urine color & appearance | 83 | 15.1 |
| Back pain | 55 | 10.0 |
| Increase in the frequency of urination | 94 | 17.2 |
| A sudden desire to go to bathroom to urinate | 87 | 15.9 |
| Fever | 33 | 6.0 |
| Abdominal pain | 67 | 12.2 |

| How much water do you drink per day? |  |  |
| --- | --- | --- |
| < 0.5 Litres | 34 | 8.8 |
| 0.5 Litres to 1 Litre | 159 | 41.2 |
| 1 Litre to 2 Litres | 142 | 36.8 |
| More than 2 Litres | 51 | 13.2 |

**Table 5:**
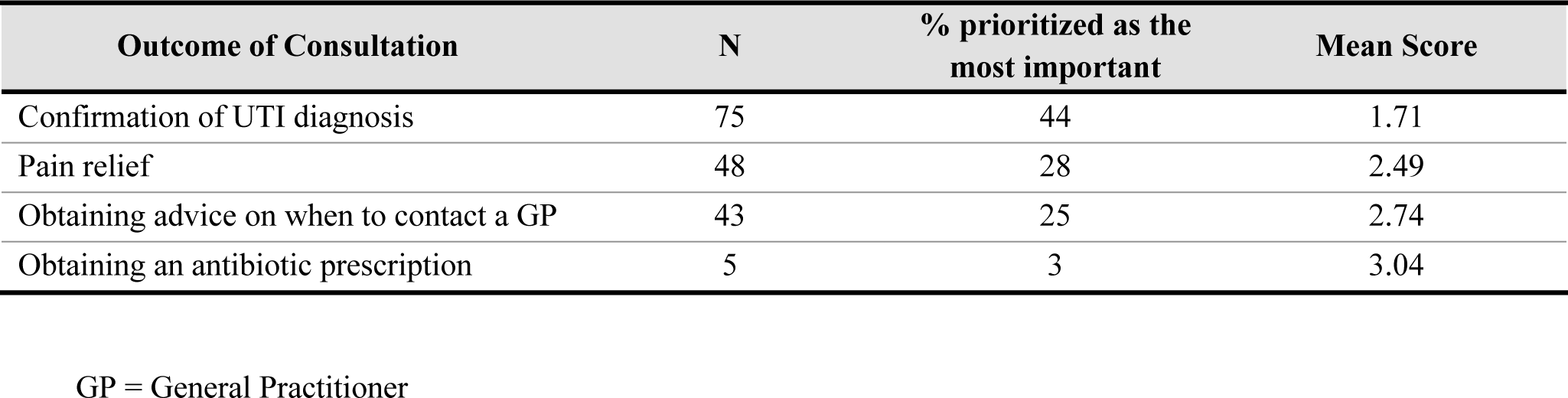
Ranking of the Outcomes of Consultation.

**Table 6:**
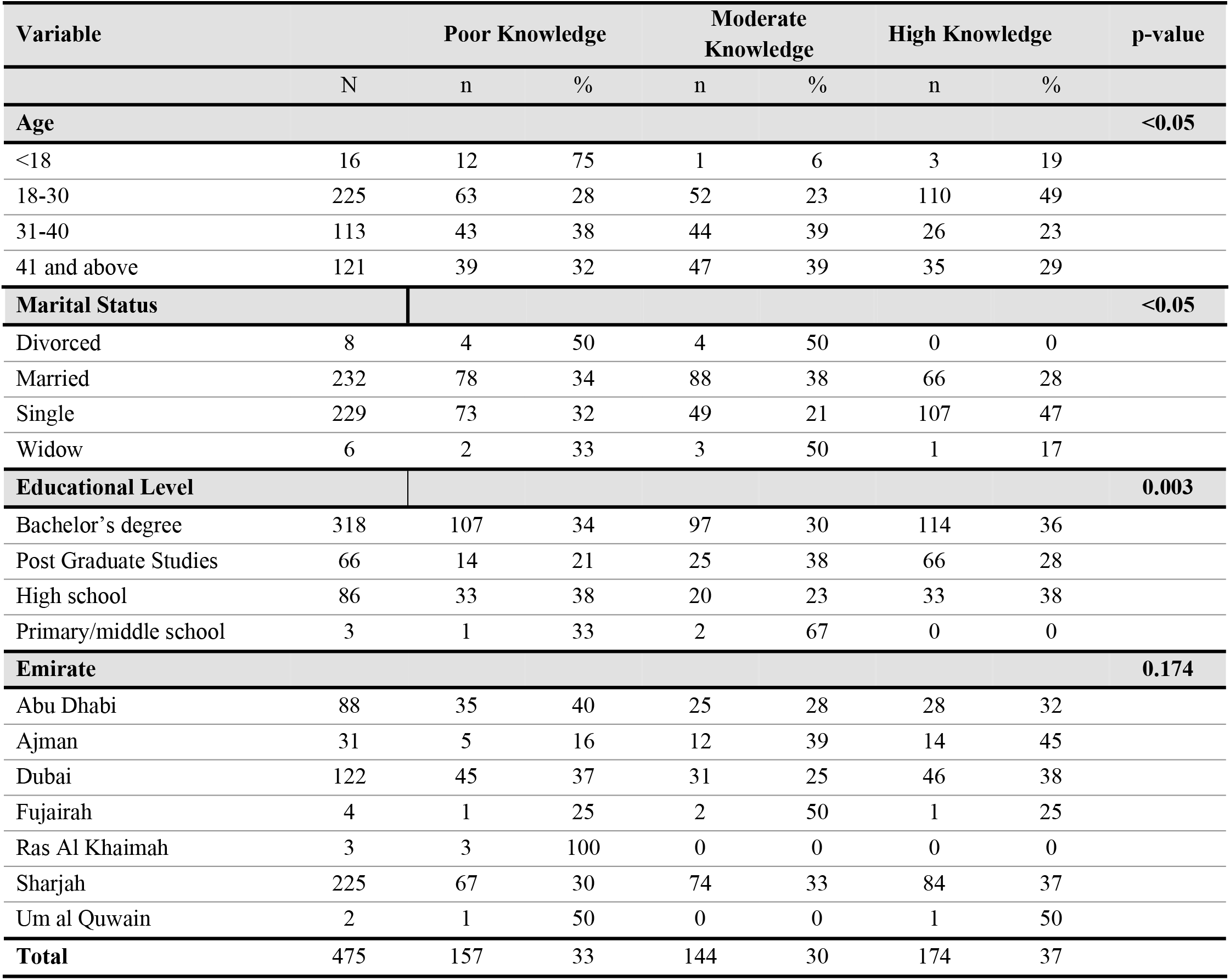
Association of Level of Knowledge of Participants Regarding UTI with Demographic Characteristics.

**Table 7:** Association of Level of Knowledge with Attitudes of Participants Regarding UTI.

| Variable | Poor Knowledge |  |  | Moderate Knowledge |  | High Knowledge |  |
| --- | --- | --- | --- | --- | --- | --- | --- |
|  | N | n | % | n | % | n | % |
| **In your opinion, what is the appropriate way of dealing with UTI?** |  |  |  |  |  |  |  |
| Drink more water | 74 | 24 | 32 | 27 | 36 | 23 | 31 |
| Go to the Hospital/Clinic | 269 | 32 | 12 | 101 | 38 | 136 | 51 |
| I don't know | 9 | 2 | 22 | 3 | 33 | 4 | 44 |
| Rest at home | 1 | 0 | 0 | 0 | 0 | 1 | 100 |
| Shower more frequently | 6 | 1 | 17 | 2 | 33 | 3 | 50 |
| Take analgesics (painkillers) | 1 | 0 | 0 | 0 | 0 | 1 | 100 |
| Take antibiotics directly | 26 | 9 | 35 | 11 | 42 | 6 | 23 |
| **Do you think UTIs are common?** |  |  |  |  |  |  |  |
| Yes | 363 | 64 | 18 | 130 | 36 | 169 | 47 |
| No | 23 | 4 | 17 | 14 | 61 | 5 | 22 |
| **Which of the following statements do you think is true regarding UTI?** |  |  |  |  |  |  |  |
| I don't know | 34 | 13 | 38 | 16 | 47 | 5 | 15 |
| UTIs affect both genders equally | 52 | 16 | 31 | 19 | 37 | 17 | 33 |
| UTIs affect females more | 295 | 39 | 13 | 104 | 35 | 152 | 52 |
| UTIs affect males more | 5 | 0 | 0 | 5 | 100 | 0 | 0 |
| **Which complications do you expect from having a UTI?** |  |  |  |  |  |  |  |
| It can damage the kidneys | 289 | 37 | 13 | 107 | 37 | 145 | 50 |
| It can decrease the quality of life | 171 | 22 | 13 | 50 | 29 | 99 | 58 |
| It can lead to recurrent UTIs | 142 | 5 | 4 | 33 | 23 | 104 | 73 |
| It can affect pregnancies | 85 | 4 | 5 | 18 | 21 | 63 | 74 |
| It can lead to death | 29 | 0 | 0 | 2 | 7 | 27 | 93 |
| It can cause weight loss | 8 | 0 | 0 | 3 | 38 | 5 | 63 |
| **Do you feel UTI is a serious condition/disease?** |  |  |  |  |  |  |  |
| I don't know | 70 | 14 | 20 | 43 | 61 | 13 | 19 |
| Yes | 178 | 23 | 13 | 42 | 24 | 113 | 63 |
| No | 138 | 31 | 3 | 59 | 5 | 48 | 4 |
| **Total** | **386** | **68** | **18** | **144** | **37** | **174** | **45** |

**Table 8:**
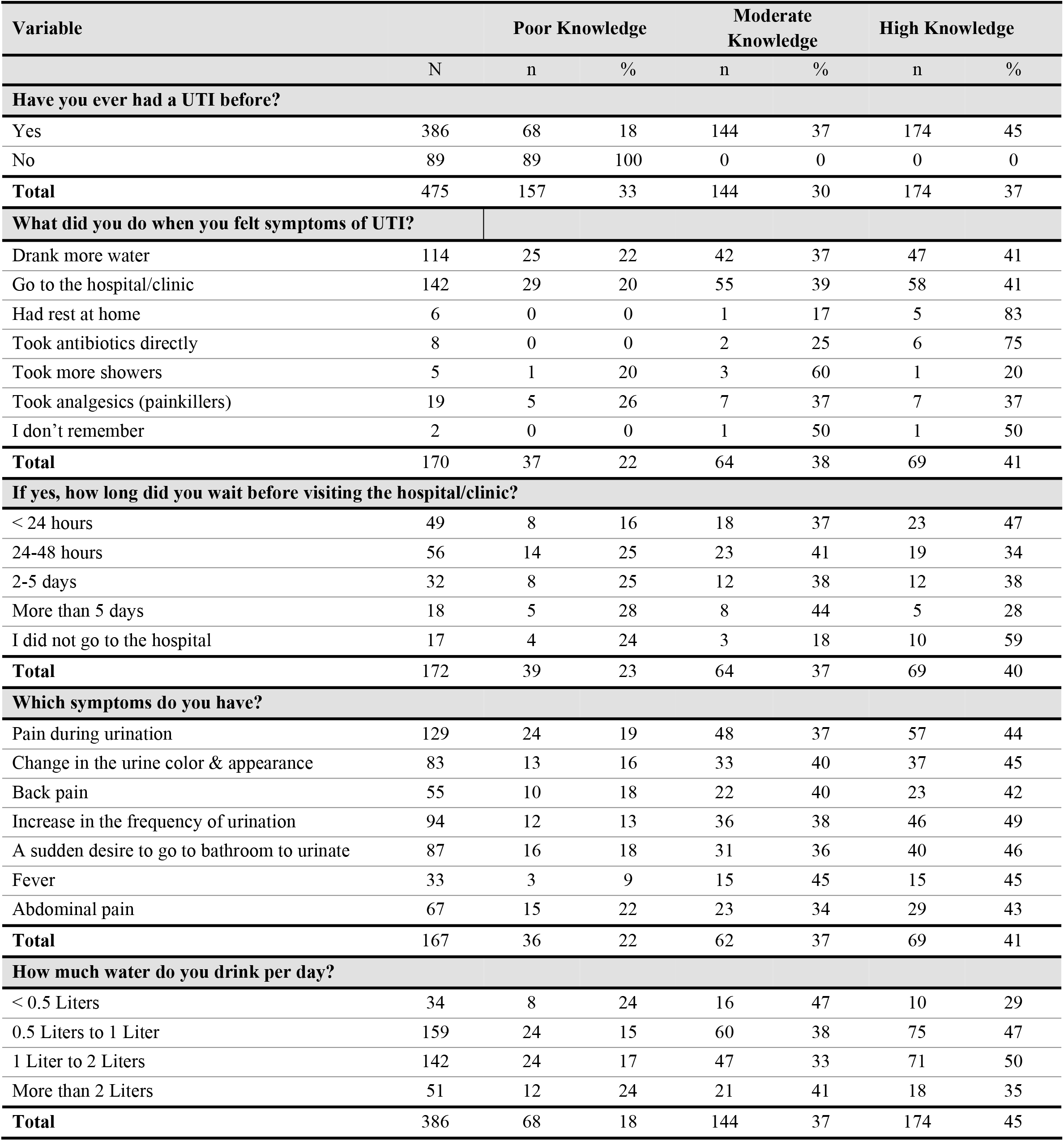
Association of Level of Knowledge with Practices of Participants Regarding UTI.

### 2. Knowledge of the Participants Regarding Urinary Tract Infections

Participants who responded ‘No’ to the question “Have you heard of the term urinary tract infection (UTI)?” were categorized as possessing a minimal level of knowledge (18.7%), and the questionnaire was subsequently concluded for them at that point. The most prevalent understanding of UTIs among these respondents is inflammation of the kidney, bladder, or urethra (53.5% chose “Can be in all the above”), followed by ‘Inflammation of the urethra’ only (23.2%). A majority (69.7%) correctly identify bacteria as the most common cause of UTIs. Additionally, most participants were able to accurately identify the symptoms, habits that increase the risk, and preventive measures of UTIs.

### 3. Attitude of the Participants Regarding Urinary Tract Infections

Most respondents (69.6%) believe that going to the hospital/clinic is the appropriate way to deal with a UTI. Drinking more water is also a common response, with 19.2% of respondents suggesting this method. 46.1% of respondents perceive UTI as a serious condition/disease. Some of the most commonly believed complications of UTIs included “damage to the kidneys” and “decrease in quality of life” (39.9% and 23.6%, respectively).

### 4. Practices of the Participants Regarding Urinary Tract Infections

With regards to practices undertaken during UTIs, among the participants with a history of UTI episodes, 56 (32.6%) waited 24-48 hours before seeking medical attention at a hospital or clinic, while 17 (10%) did not visit a hospital at all. The most common initial actions taken by these participants included visiting a hospital or clinic and increasing water intake (48% and 38.5%, respectively).

### 5. Outcomes of Consultation

Regarding participants’ expectation of consultation outcomes, participants were required to rank four possible outcomes of a visit to the GP in order of importance. The majority of the participants (44%) ranked Confirmation of UTI Diagnosis as the most important factor, with a mean ranking of 1.71. However, the least prioritized outcome was Obtaining an Antibiotic Prescription (3%), with a mean ranking of 3.04.

### 6. Association of Level of Knowledge Regarding UTI with Demographics

Distinct trends were found when comparing demographic factors with knowledge levels. Most notably, the age group of 18-30 years showed the highest percentage of high-knowledge individuals (49%) compared to other age groups, while individuals below 18 years predominantly (75%) had poor knowledge about UTIs (p = <0.05). Education level was significantly (p = 0.003) associated with UTI knowledge as Individuals with higher educational qualifications, such as a bachelor’s degree or postgraduate studies, displayed a lower percentage of poor knowledge about UTIs (34% and 21%, respectively). Moreover, the highest percentage (41%) of high-knowledge individuals was found in the postgraduate group. Marital status also exhibited a significant (p = <0.05) association with knowledge, with single individuals demonstrating high levels of knowledge (47%) compared to divorced or married counterparts. While geographical differences in knowledge levels were noted across various Emirates, these were not statistically significant (p = 0.174).

### 7. Association of Level of Knowledge with Attitudes of Participants Regarding UTI

Individuals with a high level of knowledge about UTIs were significantly (p = <0.05) associated with a greater likelihood of recognizing the seriousness of UTIs (63% of the participants who believed that UTIs are serious and were categorized as having high knowledge also) and their potential complications such as damage to the kidneys and leading to recurrent UTIs.

The belief that UTIs are more common in females was prevalent across all knowledge levels but was most pronounced (52%) in the high-knowledge group.

### 8. Association of Level of Knowledge with Practices of Participants Regarding UTI

Higher knowledge levels were associated with more proactive and appropriate health behaviors, such as seeking medical attention promptly and drinking more water. Going to the hospital/clinic was reported 41% with high knowledge but only by 20% of those with poor knowledge. Moreover, a higher proportion of individuals with high knowledge sought medical attention immediately within 24 hours (47%). Drinking more water was a common action across all knowledge levels, but more prevalent in the high-knowledge group (41%). Individuals with higher knowledge tended to drink more water, with 50% of those consuming 1-2 liters having high knowledge and 35% consuming more than 2 liters daily.

## Discussion

Urinary tract infections (UTI) are one of the most common, challenging, and misunderstood diseases that women experience as outpatients [13]. UTIs are a prevalent health issue among females globally [1], with a notable prevalence in the United Arab Emirates (UAE), which is increasing at an exponential rate [14]. The issue of delayed detection of UTIs is very prevalent. It leads to both excessive therapy and inadequate treatment [13]. If not managed properly and timely or left untreated, UTIs may cause serious complications like renal impairment, kidney scarring, sepsis, and pyelonephritis [5]. Women’s practices, attitudes, and understanding of urinary tract infections are significant variables that can affect how this illness is managed and prevented. While a substantial number of studies on this subject have been carried out globally [9-10, 13], there remains no data specific to the UAE context. This study aims to bridge this gap by exploring the knowledge, attitudes, and practices of women in the United Arab Emirates about urinary tract infections (UTIs).

Our research revealed several noteworthy findings. Most of the participants (80%) were aware of the term UTI and half of them were able to identify the correct definition. The vast majority were able to identify the most common cause of UTIs and the most prevalent symptoms. Additionally, most of them were also able to report the most appropriate ways of dealing with UTI. Therefore, the results of our study concluded that most of the respondents exhibited an acceptable level of awareness regarding UTIs. Our results regarding the knowledge of the participants are consistent with the other studies in the surrounding region like in Saudi Arabia [10, 15], and were also consistent with a study conducted in the Netherlands [9]. On the other hand, studies conducted in Bangalore [16] and Egypt [17] suggested that the participants mostly had poor knowledge and attitude, which may suggest that lower levels of knowledge are associated with developing countries. Similarly, while assessing the attitudes and practices of the respondents, we found out that a significant fraction of our study population understood the seriousness of the condition (46%) and took appropriate measures for the management of UTIs, such as seeking medical help (48%) or increasing water intake (38.5%). When asked about the outcomes of General Practitioner (GP) consultation, the participants in our study showed results similar to those of the study group in the Netherlands from Cox SML et al. [9]. In both studies, most participants chose confirmation of UTI diagnosis as the main outcome behind the GP visit, which shows that patients value getting the definitive diagnostic information during their consultation. Moreover, in our study, only 3% of the participants prioritized obtaining an antibiotic prescription compared to 14.3% in the Netherlands study [9]. These findings may be explained by the wider accessibility of antibiotics without a prescription in the UAE. Uropathogenic bacteria are notable for their frequent resistance to many drugs [18]. It is necessary to monitor the antibiotic resistance patterns in UTIs [19].

Regarding antibiotic usage, participants were asked about the appropriate way of dealing with a urinary tract infection and only 6.7% of the participants chose to take antibiotics directly, and only 2.7% took antibiotics directly when they felt UTI symptoms. This may suggest increased awareness of appropriate antibiotic use and a desire for more conservative treatment approaches in the UAE population. Moreover, this trend aligns with global health concerns regarding antibiotic resistance, signifying a positive step towards responsible antibiotic utilization within the UAE community.

Furthermore, we studied the association of knowledge level and demographics, attitudes, and practices and concluded that the individuals who possessed a high level of knowledge were 18-30 years old and were well-educated. A similar study was conducted in Malaysia, which stated a possibility that educated respondents most likely would have studied about UTI or UTI-related topics in their syllabus, which could be a possible reason for their high level of knowledge [20]. These individuals also understood the seriousness of UTIs and preferred to seek medical help for treatment. Significant associations were found between knowledge and age, educational level, and marital status; however, no significant association was found between the level of knowledge and Emirate which is in contrast to studies conducted in Saudi Arabia [10, 15].

However, certain strengths and limitations of our study need to be acknowledged. This is the first study that assessed the knowledge, attitudes, and practices of women regarding UTIs in the UAE. Enhancing patients’ understanding, beliefs, and actions about prevalent illnesses plays a crucial part in health education [21]. The results of this study can guide the tailored educational campaigns for women’s health in the UAE. The study’s limitations include the possible occurrence of recall bias due to the dependence on self-reported data, as well as the possibility of participants’ replies being influenced by social desirability or response bias.

In conclusion, our study found that most of the respondents had adequate knowledge about UTIs. The results of this study conclude that most of the women in the UAE exhibited substantial awareness and behavioral patterns regarding UTIs. Higher knowledge levels were associated with more proactive & appropriate health behaviors, such as seeking medical attention promptly & drinking more water.

## Data Availability

Not Applicable

## Acknowledgments

None to Declare

